# The impact and progression of the COVID-19 pandemic in Bulgaria in its first two years

**DOI:** 10.1101/2022.09.01.22279496

**Authors:** Antoni Rangachev, Georgi K. Marinov, Mladen Mladenov

## Abstract

After initially having low levels of SARS-CoV-2 infections for much of the year, at the end of 2020 Bulgaria experienced a major epidemic surge, which caused the highest recorded excess mortality in Europe and among the highest in the word (Excess Mortality Rate, or EMR ∼ 0.25%). Two more major waves followed in 2021, followed by another one in early 2022. In this study we analyze the temporal and spatial patterns of excess mortality at the national and local levels and across different demographic groups in Bulgaria, and compare those at the European level. The country has continued to exhibit the previous pattern of extremely high excess mortality as measured both by crude mortality metrics (EMR ∼ 1.05% up to the end of March 2022) and by standardized ones – Potential Years of Life Lost (PYLL) and Aged-Standardized Years of life lost Rate (ASYR). Unlike Western Europe, the bulk of excess mortality in Bulgaria, as well as in several other countries in Eastern Europe, occurred in the second year of the pandemic, likely related to the differences in the levels of vaccination coverage between these regions. We also observe even more extreme levels of excess mortality at the regional level and in some subpopulations (e.g. total EMR values for males ≥ 2% and EMR values for males aged 40-64 ≥ 1% in certain areas). We discuss these observations in light of the estimates of infection fatality rate (IFR) and eventual population fatality rate (PFR) made early in the course of the pandemic.

## Introduction

The SARS-CoV-2 virus and the COVID-19 disease ^1–3^ that it causes have triggered the most significant acute public health crisis in more than a century. SARS-CoV-2 has spread widely in most countries around the world, and has been the driver of substantial excess mortality in many of them ^4,5^.

The pandemic took divergent trajectories in different regions of the world, initially depending on the timing of the imposition of containment measures relative to the undetected early cryptic spread of the virus, and later based on some combination of the relaxation of these measures, seasonal effects, the build up/waning of population immunity, the appearance of new variants of SARS-CoV-2 that are more contagious and/or antigenically divergent, and other factors. Some countries were heavily affected early on and then experienced further major epidemic waves, others were only hard hit at later stages of the pandemic.

By the end of 2020, Bulgaria emerged as one of the countries experiencing among the highest pandemic-related excess mortality in the world, even though it was one of the early containment success stories in the course of the pandemic, largely escaping the first major wave that affected greatly many areas in Western Europe, and the Americas. As a previous analysis of ours has shown ^6^, the EMR value for the country by January 1st 2021 stood at ∼ 0.25% (more than twice the official death count, due to some combination of insufficient testing, registration of COVID deaths as having occurred due to other reasons, and elevated mortality from otherwise treatable other conditions due to hospital capacity being exceeded).

Subsequently, the country experienced three more major waves, in March-April 2021, in the last few months of 2021, and early in 2022. In this study we track the development and assess the impact of the pandemic on different demographic groups and regions in Bulgaria up to the end of March 2022, using a combination of excess mortality analyses and SARS-CoV-2 genome sequencing surveillance.

These subsequent waves have dramatically increased the excess mortality burden in the country, and as a result it has become the first one (among those for which overall mortality data is available) where COVID-related excess deaths have exceeded 1% of the total population. Furthermore, we, continuing the trend established previously ^6^, observe major discrepancies between the outcomes within the country. EMR values in some regions are now approaching 2%, and they have exceeded that value for males in certain areas. In addition, mortality in the working age 40-64 group is approaching or has even exceeded 1%, a surprising result considering the commonly assumed dramatic age skew of COVID-related mortality. Despite the reduced Case Fatality Ratio (CFR) associated with the newly emerged at the end of 2021 Omicron variant, considerable excess mortality, not captured by official COVID death statistics, persisted in the first months of 2022. These patterns are in stark contrast to those observed in countries in Western Europe, where excess mortality was concentrated in 2020 and decreased in 2021. They are, however, shared with most other countries in Eastern Europe, although Bulgaria still exhibits the most extreme excess mortality figures. The likely explanation for this pattern is the lower vaccination rates in Eastern Europe and particularly in Bulgaria. Finally, we discuss these findings in the context of the commonly cited figures for the infection fatality rate (IFR) of COVID-19.

## Results

### Loss of life as a result of the COVID-19 pandemic

In order to evaluate the impact of the COVID-19 pandemic on different countries in Europe we applied excess mortality analysis for the period from the start of the pandemic until the end of March 2022, following previously established methods ^4,6^ (see the Methods section for details). Excess mortality measures are more objective measures of pandemic impact as officially recorded COVID mortality is often not an accurate representation of reality, due to insufficient availability of testing, inaccurate reporting, and other factors, such as second-order impacts of COVID infections (i.e. overwhelmed healthcare systems not being able to provide adequate treatment) leading to fatalities that would not occur under normal circumstances. Specifically in Bulgaria, 95% of the officially confirmed COVID-19 deaths occurred in hospitals, meanng that few of those who died outside hospitals entered official statistics. The view that most excess deaths are due to COVID-19 is supported by the observation that the trajectory of excess deaths generally closely tracks that of officially recorded COVID-19 cases and deaths. Considerable discrepancies can be observed between official statistics and excess deaths, with excess deaths exceeding official numbers by even an order of magnitude or more in multiple countries ^4^, underscoring the importance of analyzing excess mortality to accurately understand the real impact of the pandemic. During its first major wave in 2020, Bulgaria exhibited not only the highest excess mortality in the European Union, but also one of the highest discrepancies between excess deaths and official COVID deaths, with an “undercount ratio” of 2.52 × ^4,6^.

We previously estimated that Bulgaria had lost 19,004 lives during its first major COVID wave in 2020. The updated analysis up to the end of March 2022 reveals that this number has increased to 68,569 (95% CI: ± 6,772), compared to an official COVID death count of 36,529 ^7^, i.e. the current undercount ratio is 1.88 × (± 0.18). In 2021, results from the most recent nationwide census for Bulgaria became available, which showed a decrease of the population down to 6,520,314 ^8^. Accounting for this updated denominator estimate, the EMR value for Bulgaria has now exceeded 1%, standing at 1.05% circa March 31 2022. This is the highest value recorded in any country for which excess mortality data is available ^4^.

As crude mortality measures such as the EMR and the P-score (the percentage increase in mortality relative to baseline) may not be optimal for comparisons between populations with different demographic structures, we also calculated two standardized measures that control for such variation and aim at measuring the years of life lost as a result of the pandemic: the Potential Years of Life Lost (PYLL) and Aged-Standardized Years of life lost Rate (ASYR; see the Methods section for details). Figure 1 shows standardized (per 100,000 population) ASYR and PYLL values for European countries in the three years of the pandemic, in total and for males and females separately. Bulgaria exhibits the highest mortality by all measures among this set of countries (per-100,000 PYLL values of 12,370, 10,983 and 13,002 and ASYR values of 11,516, 9,157 and 13,745 in total, and for females and males, respectively), followed by Lithuania and Poland. Excess mortality in Eastern Europe countries is much higher than that in Western Europe, and, curiously, is concentrated in the year 2021 rather than 2020, while the opposite pattern is observed in severely affected early in 2020 countries such as Spain and Italy. This observation is likely explained by two factors. First, the pandemic in 2021 in Europe was dominated first by the Alpha ^10^ and then by the Delta ^11^ SARS-CoV-2 variants, which are known to cause more severe disease than the ancestral wild-type (WT/D614G) virus ^10,12^. Second, COVID-19 vaccination rates in Eastern Europe have been consistently lower than those in Western Europe (for example, only 11.5% of the population in Bulgaria had received two vaccine doses by July 1st 2021, and this number only increased to 29.6% by the end of March 2022 ^7^), meaning that the Alpha, and especially the Delta waves encountered a much larger proportion of completely immunologically naive individuals in populations in Eastern Europe than in Western Europe, resulting in the observed disproportionally higher mortality in the former. Indeed, we find strong inverse correlation between vaccination rates and excess mortality, in particular in 2021 (Pearson *R*^2^ = 0.57, *p* ≤ 0.0001, and Spearman *r* = − 0.69, *p* ≤ 0.0001 for ASYR values, and Pearson *R*^2^ = 0.56, *p* ≤ 0.0001; Spearman *r* = − 0.65, *p* = 0.0001 for PYLL; Supplementary Figure 1).

**Figure 1:**
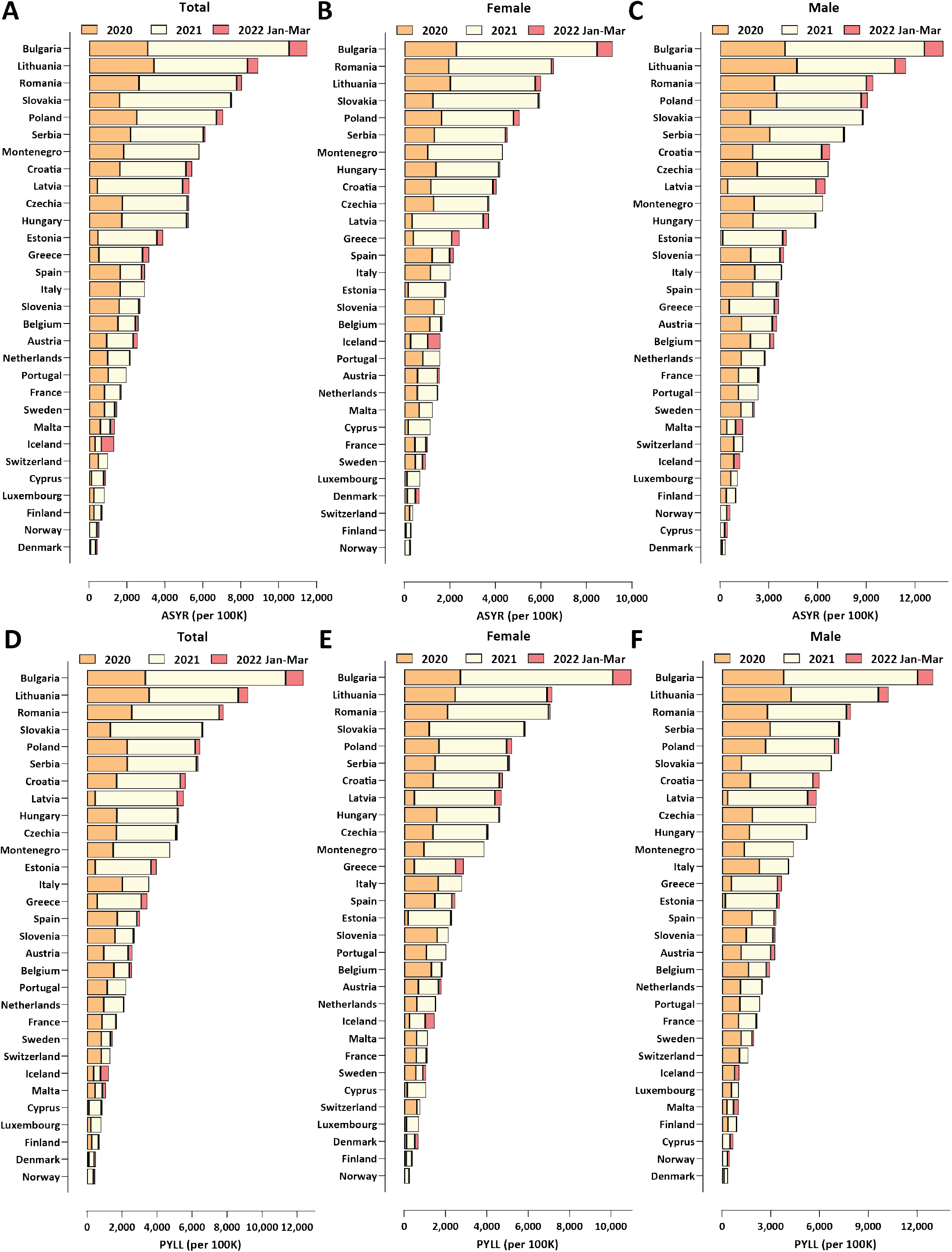
Excess mortality in Europe and Bulgaria during the COVID-19 pandemic (up to the end of March 2022). (A) Standardized ASYR values, total; (B) Standardized ASYR values, females; (C) Standardized ASYR values, males; (D) Standardized PYLL values, total; (E) Standardized PYLL values, females; (F) Standardized PYLL values, males.

We estimate that each excess death in Bulgaria resulted in 11.70, 12.70, and 10.43 years of life lost overall, for males, and for females, respectively, based on the ASYR metric, and in 12.57, 12.02, and 12.51 years of life lost overall, for males, and for females, respectively, based on the PYLL metric (Supplementary Figure 2).

Finally, we observe that male mortality is consistently higher than that of females for all the countries examined, consistent with previous observations ^16^.

### Temporal trajectory of the pandemic in Bulgaria

Figure 2 shows the evolution of the SARS-CoV-2 variant composition in Bulgaria based on available genome sequencing data ^17^. The first major wave, in late 2020, was driven by WT-like (i.e. with the addition of the D614G mutation ^18–20^ but otherwise without major spike protein mutations affecting antigenic properties) B.1.x lineages. The Alpha variant came to dominate in early 2021 and drove the second wave, and was then itself replaced by the Delta variant in June-July 2021. Finally, in early 2022, the Omicron BA.1 variant ^21,22^ displaced Delta and triggered the fourth major wave, with the Omicron BA.2 lineage ^23,24^ beginning the next variant displacement cycle at the end of the observation period.

**Figure 2:**
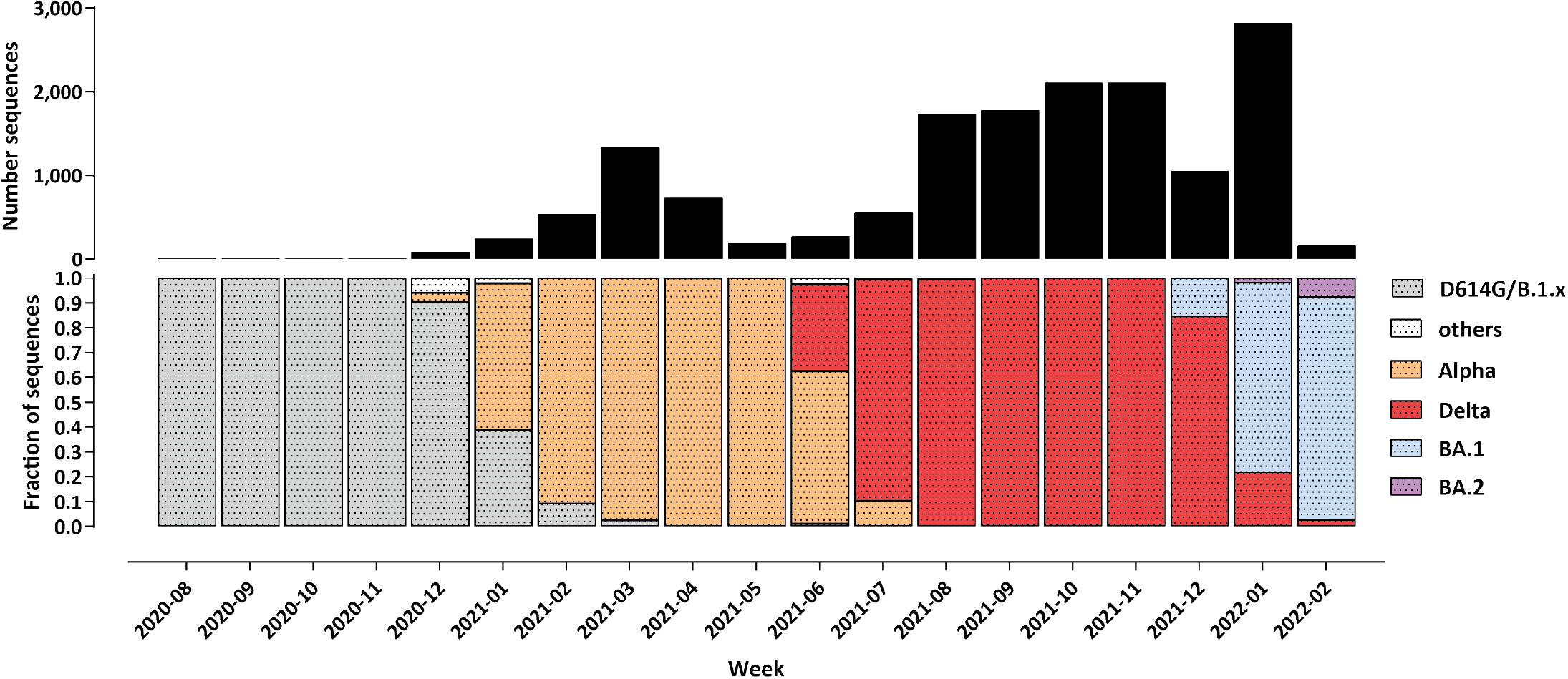
Evolution of the SARS-CoV-2 variant composition in the course of the COVID-19 epidemic in Bulgaria up to March 2022. Shown is the fraction of sequenced genomes belonging to each of the listed variants for each month since the beginning of December 2020. The total number of sequenced SARS-CoV-2 genomes is shown on top.

Figure 3 shows the trajectory of the pandemic in Bulgaria in terms of recorded clinical impacts and excess mortality. We estimate that the first wave caused ∼ 19,000 excess deaths (or EMR ∼ 0.29% of the population), the Alpha wave had a slightly lower peak and caused ∼ 15,000 (EMR ∼ 0.23%), the Delta wave peaked at about the same heights as Alpha, but was much more prolonged (Figure 3A-B), and thus caused the highest number of excess deaths – ∼ 28,000 (EMR ∼ 0.43). The largest number of infections were recorded during the Omicron wave (Figure 3A) but it caused the fewest excess deaths, at ∼ 7,000 (EMR ∼ 0.11%). A similar pattern is observed in the evolution of case fatality rate over time, which decreased dramatically once Omicron came to dominate (Figure 3C), consistent with worldwide observations of lower disease severity with the BA.1 variant than with preceding non-Omicron ones ^25,26^.

**Figure 3:**
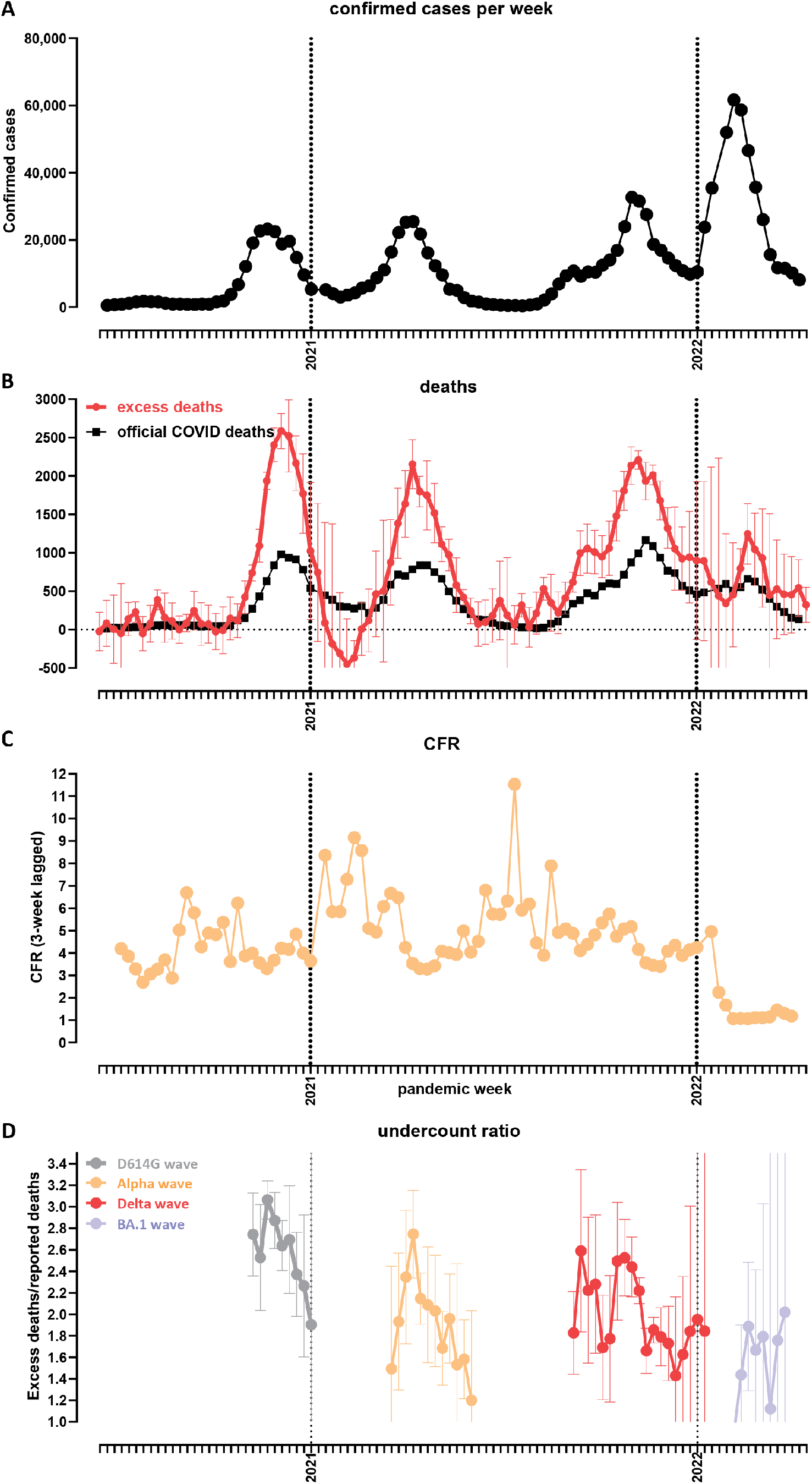
Temporal trajectory of the COVID-19 pandemic in Bulgaria. (A) Confirmed cases over time (weekly); (B) Officially reported and excess deaths over time (weekly); (C) CFR over time. Data on reported cases and deaths was obtained from the Our World In Data website ^7^; (D) Evolution of the undercount ratio (excess mortality divided by official COVID deaths) over time (note that the periods between waves, for which estimates of excess mortality are uncertain, are omitted from the graph).

Finally, we examined the “undercount ratio” (i.e. the ratio between excess deaths and official COVID deaths). Its values were highest, in the 2.5–3 × range, during the first major wave, then decreased to the 1.5–2.4 × range during the Alpha and Delta waves, and further decreased to ∼ 1.5 × during Omicron (Figure 3D). The most likely in our view interpretation of these patterns is that the undercount ratio is dependent on the extent to which hospital systems are overwhelmed by surges of severe COVID-19 cases; thus the Omicron wave, which caused the fewest excess deaths, was most accurately captured in official statistics, as proportionally fewer people died outside of the hospital system, which was able to accomodate a larger share of the severe cases than in previous waves. However, even with Omicron, large unaccounted for excess mortality still persisted, likely due to the aforementioned issues of lack of testing and improper recording of causes of death.

### Regional mortality patterns in Bulgaria

Next, we mapped the regional patterns of excess mortality in Bulgaria (Figures 4, 5 and 6 and Supplementary Figure 3). Previously ^6^, we identified a stark difference between major population centers, especially the capital Sofia, and the peripheral provinces, explained by the unfavorable demographic structure and socioeconomic characteristics of the latter (where the long-term trend has been towards depopulation, resulting in a very high median age, and an attendant decline in the availability of healthcare resources). This pattern has continued in the next three waves, and thus Sofia (city) still exhibits the lowest excess mortality in Bulgaria (EMR = 0.67%; Figure 4A-B). In contrast, excess mortality has reached as high as 1.8% in Vidin, 1.55% in Montana, and 1.5% Razgrad. Overall excess mortality is below 1% in only five Bulgarian regions, with the northeast and northwest regions showing the highest values.

**Figure 4:**
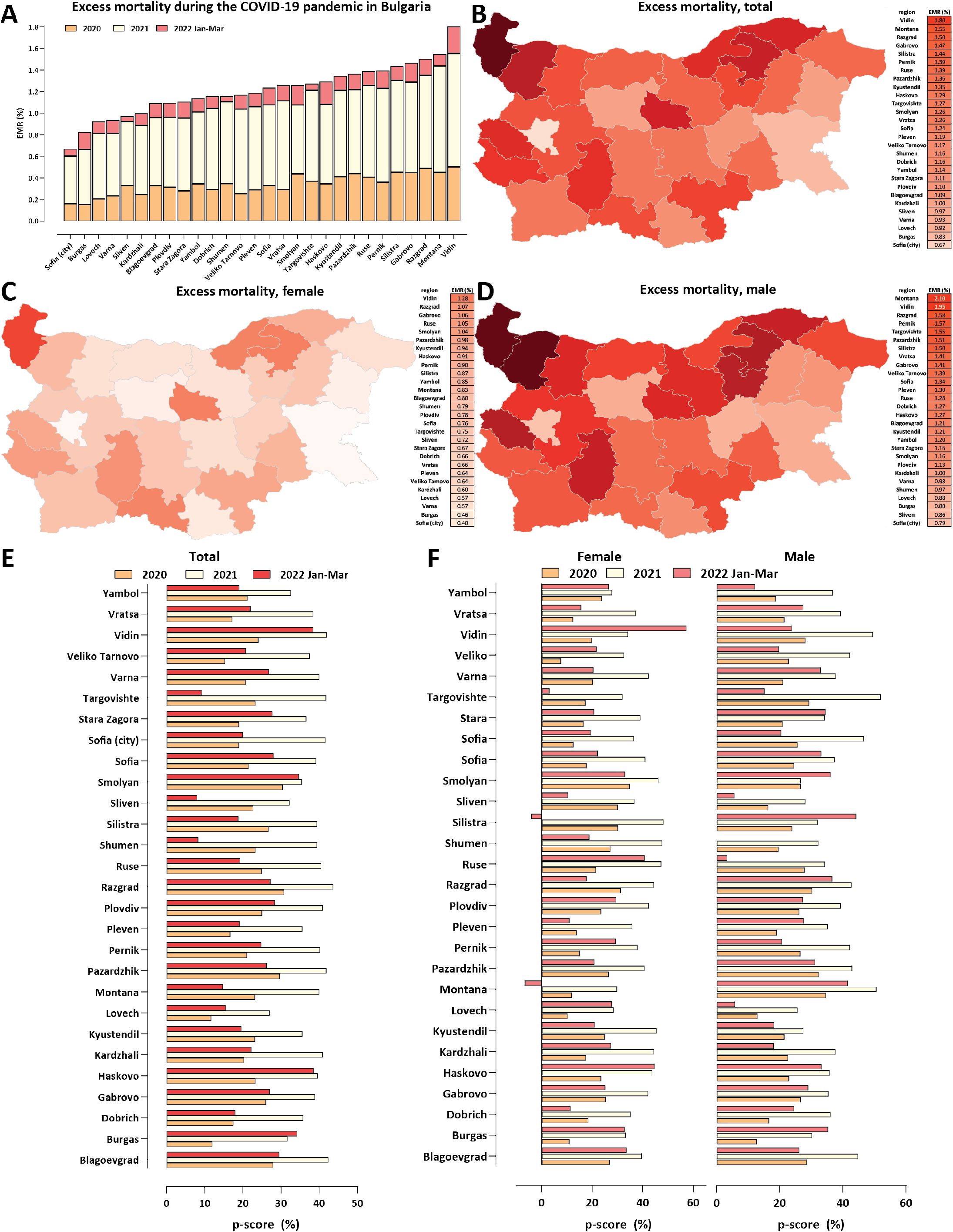
Regional excess mortality patterns in Bulgaria during the COVID-19 pandemic (up to the end of March 2022). (A) Excess and official mortality by region and year; (B) Excess mortality by region, total; (C) Excess mortality by region, males; (D) Bulgaria, Excess mortality by region, females; (E) Excess mortality (P-scores) per year; (F) Excess mortality (P-scores) for males and females per year.

**Figure 5:**
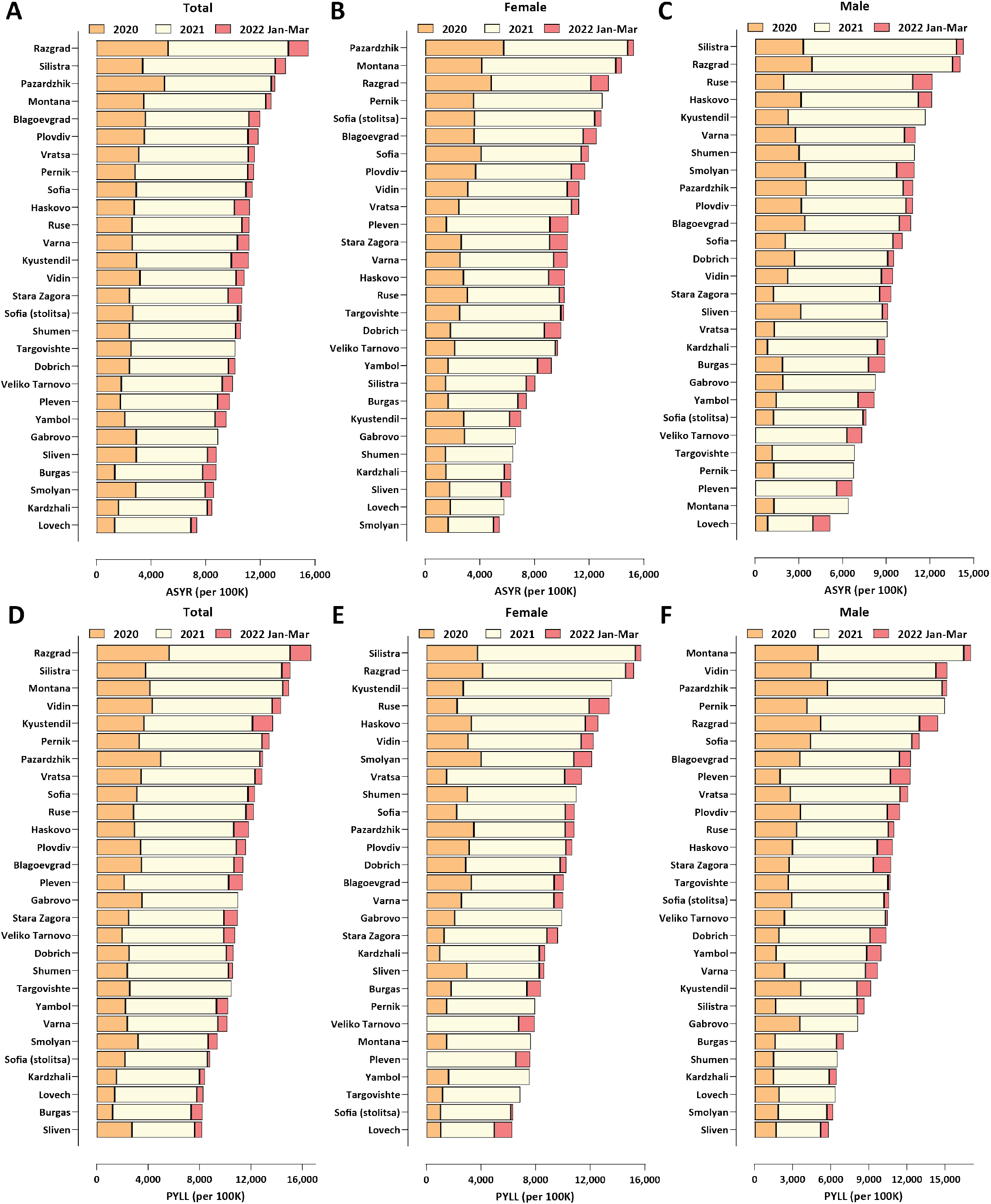
Regional excess mortality patterns in Bulgaria during the COVID-19 pandemic (up to the end of March 2022). (A) Total, standardized ASYR values; (B) Female, standardized ASYR values; (C) Male, standardized ASYR values; (D) Total, standardized PYLL values; (E) Female, standardized PYLL values; (F) Male, standardized PYLL values.

**Figure 6:**
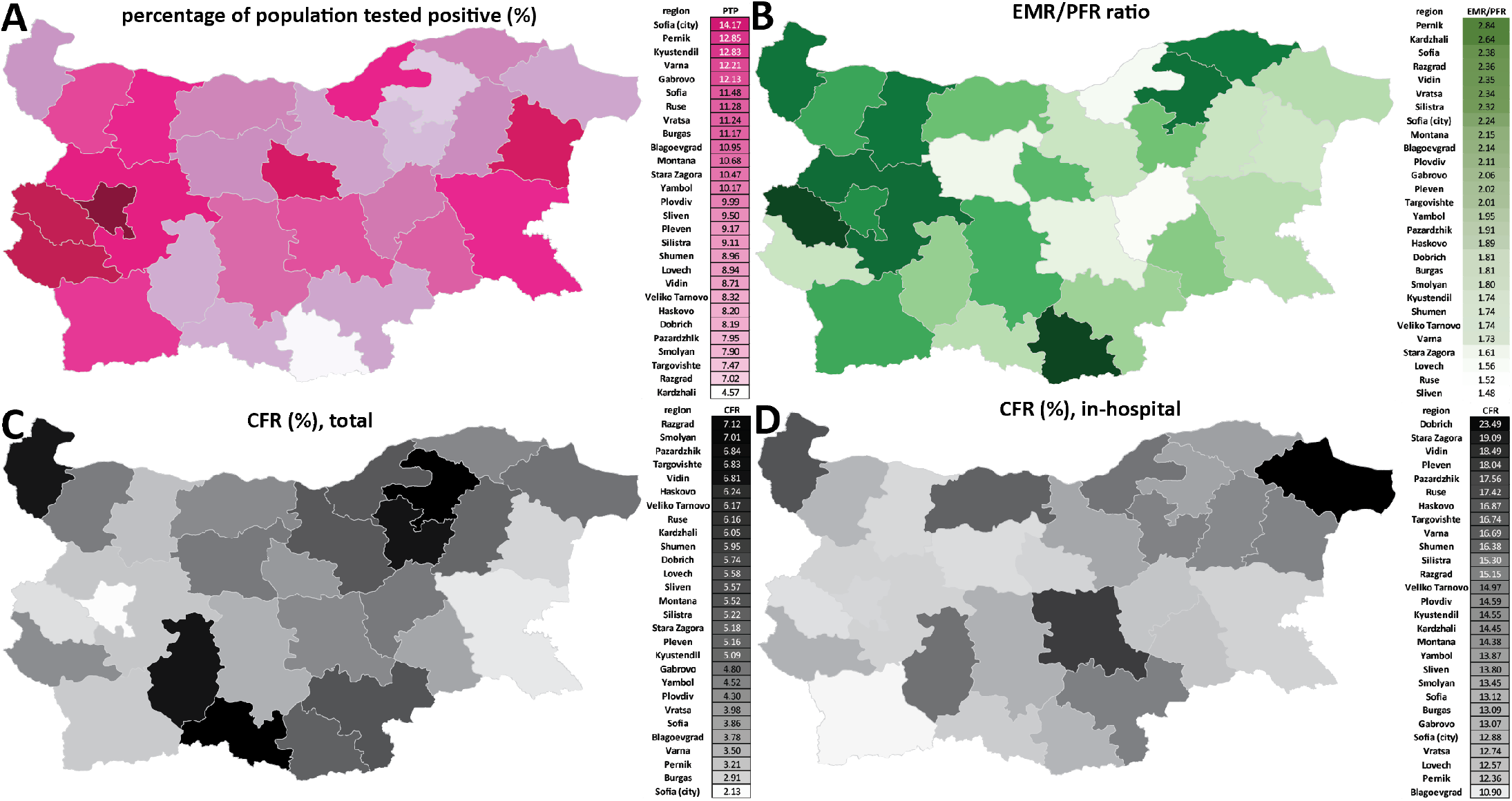
Regional discrepancies in the extent of recording of the impact of the COVID-19 and hospital outcomes in Bulgaria. (A) Percentage of the population who have tested positive for SARS-CoV-2 in Bulgarian regions; (B) Undercount ratio for Bulgarian regions; (C) Total CFR values for Bulgarian regions; (D) In-hospital CFR values for Bulgarian regions.

We observe even more extreme values for sex-specific excess mortality (Figure 4C-D) – male EMR is 2.1% in Montana and 1.95% in Vidin. Female-specific excess mortality is considerably lower in all regions, with only five of them exceeding EMR = 1% (it is highest in Vidin at 1.28%).

We also examined excess mortality using the P-score metric for each year of the pandemic (Figure 4E-F). This analysis confirmed the previously discussed observation of very high excess mortality centered on the year 2021, but also showed that in most regions excess mortality in the first quarter of 2022 has been comparable to that in 2020, despite the less severe phenotype of the Omicron variant. This observation is explained by the successful containment measures in the first half of 2020, contrasting with the very large number of infections in 2022.

We also analyzed regional excess mortality using the standardized ASYR and PYLL metrics (Figure 5). These comparisons revealed a somewhat different picture than crude mortality comparisons – ASYR and PYLL values are not lowest in Sofia city, and according to ASYR and PYLL metrics the northeastern provinces of Razgrad and Silistra have been more heavily affected than the northwestern ones of Vidin and Montana. This is likely because of the more extreme age skew of the demographic structure of the latter, which is normalized for by the ASYR and PYLL metrics but not by crude EMR estimates. As with EMR metrics, even more extreme values are observed than the already very high one for Bulgaria as a whole – e.g. in Razgrad region both ASYR and PYLL values approach 16,000 per 100,000 population.

Considerable region discrepancies are also present regarding the documenting of the pandemic and the hospital outcomes for COVID-19 patients. Unfortunately, no serological survey (of any kind, not just the anti-nucleocapside protein ones that could distinguish evidence for previous infections from vaccination with mRNA or adenoviral vaccines that target only the spike protein) has ever been carried out in Bulgaria, but it is highly likely that towards the end of March 2022, a majority of the population has been infected by SARS-CoV-2 (given the observed excess mortality; to be discussed further below). However, the percentage of the population who have tested positive is highest in Sofia city, at only 14.17%, and is as low as 4.57% in the peripheral Kardzhali region (Figure 6A). Thus, testing has been highly inadequate throughout the pandemic, with most infections remaining undocumented.

The undercount ratio between the EMR and the officially documented population fatality rate (PFR) ranges from 1.48 × in Sliven region to 2.84 × in Pernik (Figure 6B). The overall CFR ranges from 2.13% in Sofia city to ≥ 7% in Razgrad and Smolyan (Figure 6C). These discrepancies are in large part due to the inadequate testing in some of the peripheral regions in the country, which also tend to be the ones with the lowest percentage of the population that has tested positive.

Remarkably, when focusing on the CFR for hospitalized patients specifically, we find no region in which fewer than 10% of COVID-19 patients died, and in Dobrich region the number exceeds 23% (Figure 6D), underscoring the unequal and inadequate access to high-quality COVID-19 treatment across the country.

### COVID-19-related working-age excess mortality in Bulgaria and Europe

Finally, we mapped the regional patterns of excess mortality for working-age populations (Figures 7 and 8). We focused on the age 40-64 subpopulation as COVID-19-related deaths and excess mortality are low in absolute number in the younger demographics, resulting in statistically unreliable estimates at the regional level.

**Figure 7:**
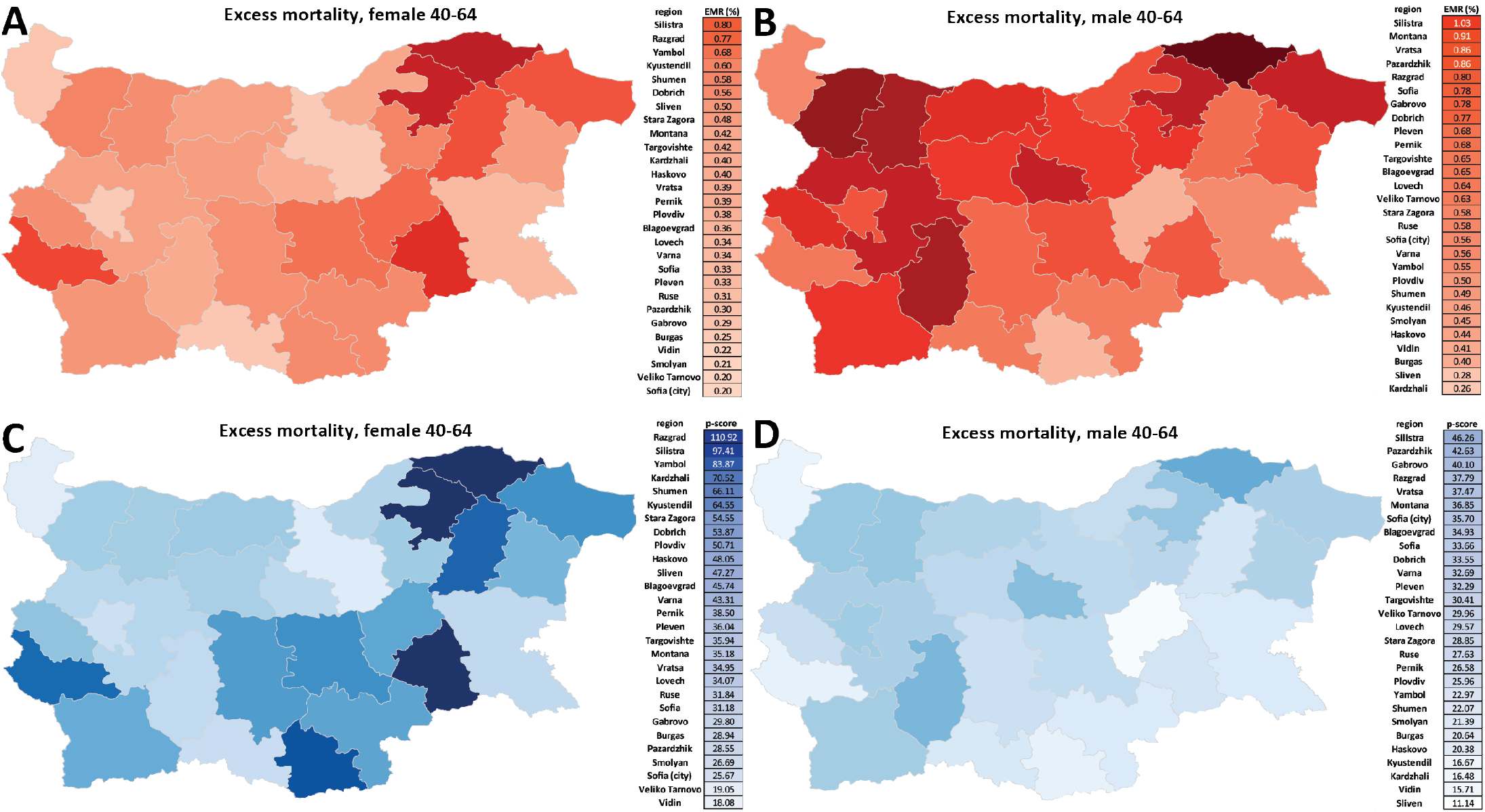
Working-age excess mortality in Bulgaria during the COVID-19 pandemic (up to the end of March 2022). (A) Excess mortality by region, females ages 40-64, EMR values; (B) Excess mortality by region, males ages 40-64, EMR values; (C) Excess mortality by region, females ages 40-64, P-scores; (D) Excess mortality by region, males ages 40-64, P-scores.

**Figure 8:**
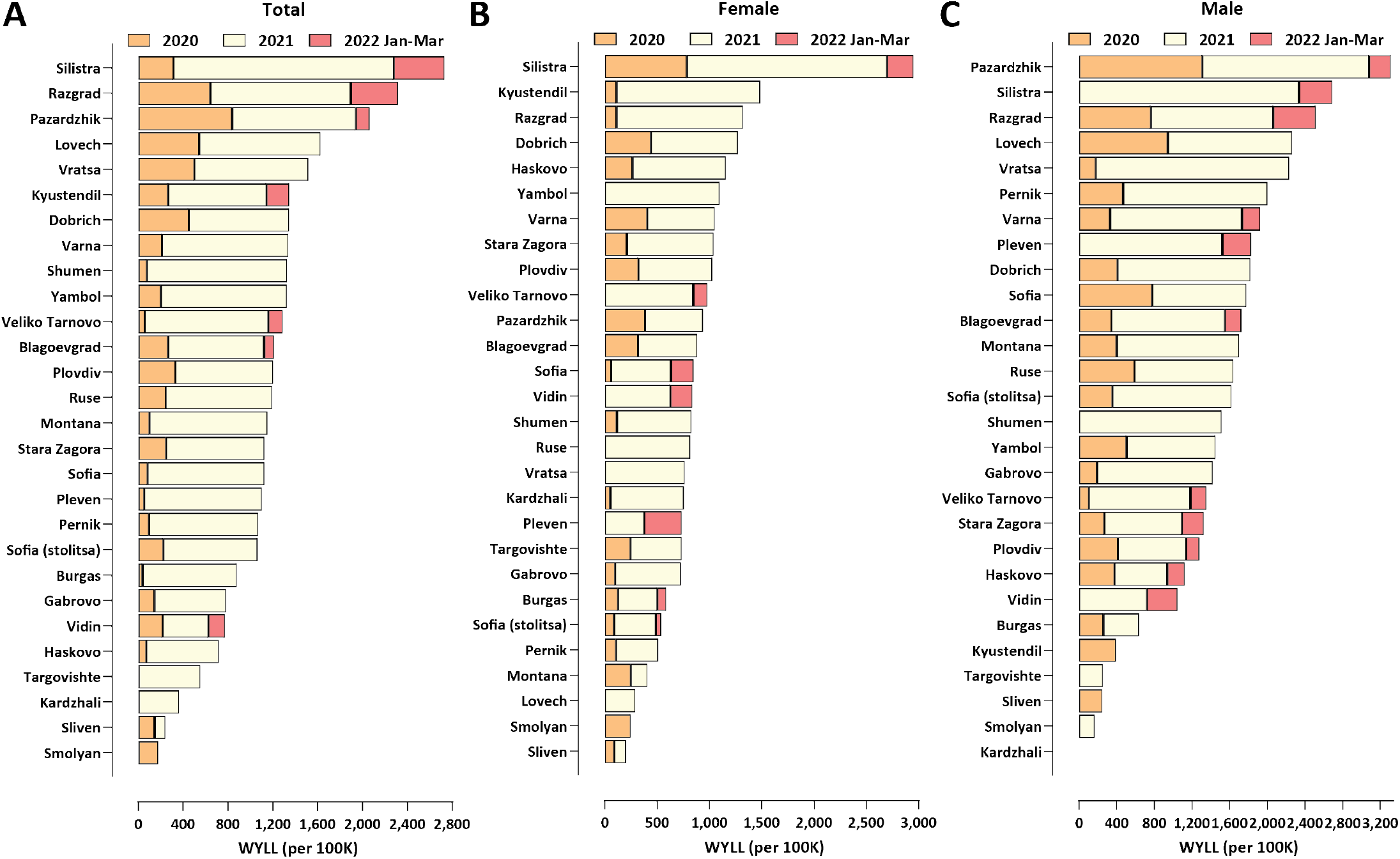
Working-age excess mortality in Bulgaria during the COVID-19 pandemic (up to the end of March 2022). (A) Total, standardized WYLL values; (B) Female, standardized WYLL values; (C) Male, standardized WYLL values.

In total, excess deaths in the 40-64 group in Bulgaria amount to 11,986 (95% CI: ± 693). We find that EMR values for this group exceed 0.2% in all regions even for females, and reach as high as 0.8% for females and 1.03% for males in Silistra region (Figure 7A-B). Working-age excess mortality has been concentrated in the northeastern and southern regions of the country (Figure 7C-D).

We also applied a standardized analysis using the Working Years of Life Lost (WYLL) metric (see the Methods section for details), which largely confirmed these regional patterns (Figure 8) – in Silistra region the WYLL value exceeds 2,500 per 100,000 population, followed by Razgrad and Pazardzhik. We also note that a unique feature of regions such as Razgrad and Silitra is the very high female-specific WYLL, at nearly double that observed in other areas, and also doubling the normal death rate (p-scores nearly or exceeeding 100%).

The average working years of life lost per excess death are 8.26 for Bulgaria as a whole, and 8.18 and 8.87 for females and males, respectively.

Finally, we compared working-age excess mortality across European countries (Figure 9). Bulgaria stands out in this analysis as exhibiting standardized WYLL values far in excess of those in other countries included in the comparison (≥70% higher than the next ranked country, Romania). As in the comparison of overall excess mortality, countries in Eastern Europe exhibit considerably higher working-age excess mortality than those in Western Europe, and it is concentrated in the second year of the pandemic.

**Figure 9:**
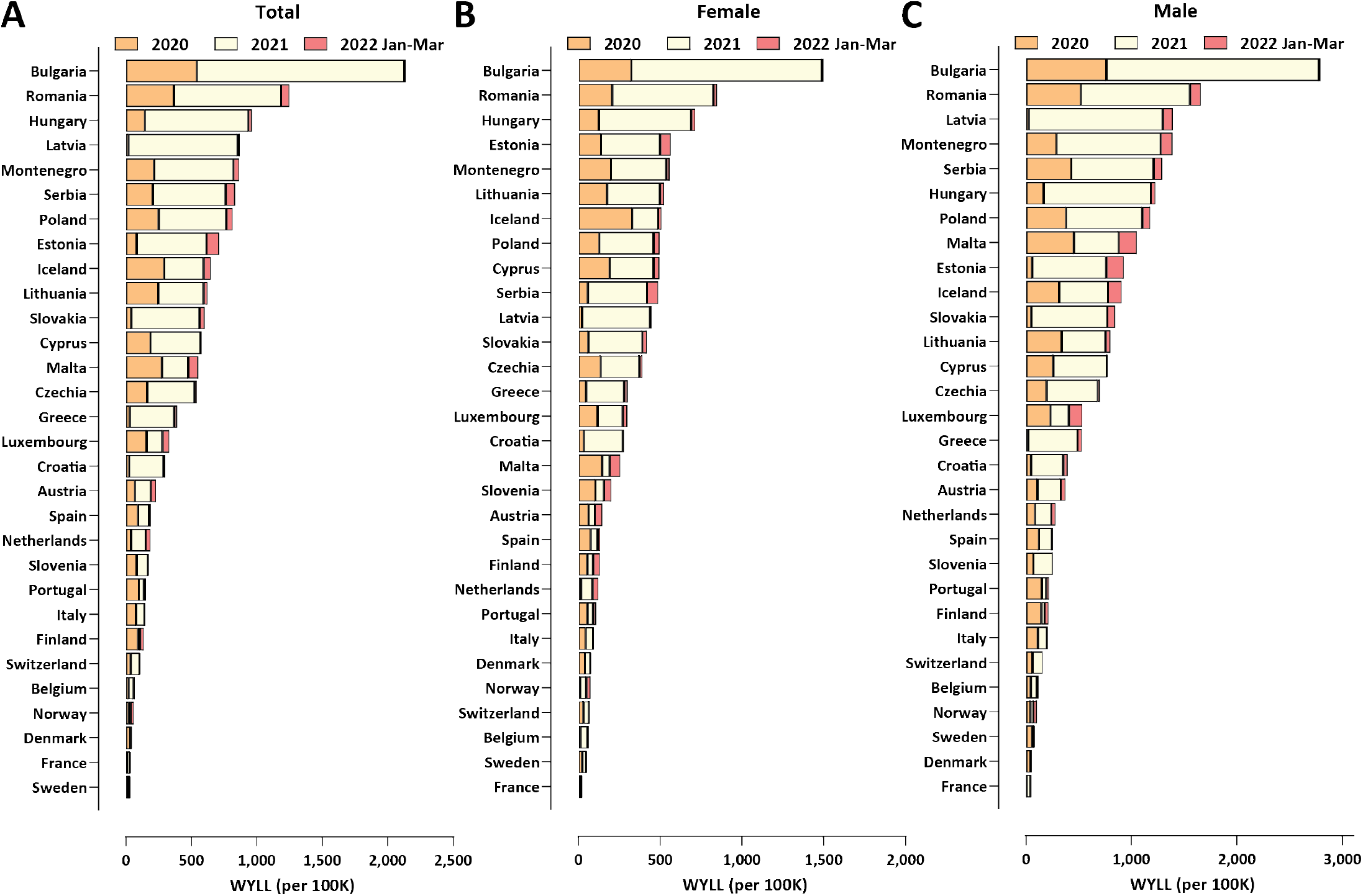
Working-age excess mortality in Europian countries during the COVID-19 pandemic (up to the end of March 2022). (A) Total, standardized WYLL values; (B) Female, standardized WYLL values; (C) Male, standardized WYLL values.

## Discussion

In this study, we map out the patterns of COVID-19-related excess mortality in Bulgaria across time, space and different demographic groups. Three striking observations stand out in the available data.

First, considerable discrepancies exist in the impact of the pandemic at the regional level, with peripheral areas of the country exhibiting much higher absolute excess mortality than the capital Sofia, presumably due to the better access to healthcare resources and the more favorable demographic structure in the latter, and possibly also the less favorable health status of the population in the former. Cardiovascular disease (CVD) is a well-known risk factor for severe COVID-19 outcomes so we examined the correlation between the CVD burden in different Bulgarian regions and excess mortality during the pandemic (Supplementary Figures 4 and 5). We find a strong positive correlation (Pearson *R*^2^ = 0.4, Spearman *r* = 0.59) for overall excess mortality and CVD burden, and weaker such correlation (Pearson *R*^2^ = 0.17, Spearman *r* = 0.43) for male-specific age (40-64) excess mortality; these observations support such a link as one of the contributing factors (we note that we also find no such correlation for female-specific working age excess mortality and for the standardized ASYR and PYLL metrics; this is likely because CVD disease burden manifests itself earlier in males and because ASYR and PYLL place less weight on excess mortality in the very elderly where CVD burden is most pronounced). The other likely major contributing factor to regional discrepancies is the unequal distribution of healthcare resources, as we previously discussed in more detail ^6^.

Second, overall excess mortality in Bulgaria is extremely high, as it is now well in excess of 1% of the total population.

This result is very important for the overall understanding of the pandemic as it finally places the early estimates of the potential impact of the SARS-CoV-2 virus in a proper context.

Numerous estimates for SARS-CoV-2’s IFR have been published, particularly early in the pandemic. A major survey of available data ^12^ estimated the age-standardized IFR for Bulgaria to be 0.873% in early 2020, decreasing to 0.565% in early 2021 (likely thanks to improved treatments). An early-2020 estimate for Belgium ^27^ placed the overall IFR at ∼ 1.5%. Published early-2020 estimates for Spain were for IFR = 1.2% ^28^ and 1.15% ^29^. For Eastern Europe as whole, an IFR value ∼ 1.45% has been published ^30^. Other estimates include 0.6% for the early-pandemic IFR in China ^31^, 0.5% in Switzerland and 1.4% in Lombardy, Italy ^32^ during the early-2020 wave, and meta-analysis-based overall estimates of 0.68% ^33^ and 1–1.5% ^34^.

In addition, several much lower values were also published during the first year of the pandemic, such as IFR at 0.04% ^35^, a global one at ∼ 0.15% ^36^, IFR at 0.17% ^37^ for Santa Clara County in California, and others.

The validity of these estimates can be evaluated in the light of the fact that Bulgaria’s excess mortality stood at 1.05% in March 2022, and that in some regions of the country it approached 2%. This outcome is the result of a combination of the following factors. First, a majority of the population must have been infected by that point (otherwise the IFR in Bulgaria would have to exceed 2%, which is unlikely), although how many exactly have been infected is not possible to say in the absence of an anti-nucleocapside serosurvey (and even then, seroreversion would probaly bias estimates downwards). Second, reinfections became an increasingly common phenomenon, first with the arrival of the Delta variant ^38^, and especially after the appearance of Omicron. Third, the virulence of SARS-CoV-2 prior to Omicron was increasing, with the Alpha variant being more severe than the WT and the Delta variant being even more severe than Alpha; meanwhile the IFR estimates from 2020 and early 2021 were based on the WT virus. Finally, vaccination in Bulgaria remained very low throughout the examined period, meaning that the Delta and Omicron waves were met with a large population of immunologically naive individuals, resulting in much higher mortality than in countries with high vaccination coverage. While deeply regrettable as a public health outcome for the country, this fact allows the observation of the potential full impact of the pandemic after infecting most of a population with a high median age and in the absence of vaccination, a situation that has been avoided, at least for the time being, in most other countries with similar demographic structures.

Third, we also observe extremely high excess mortality in working age populations, far higher than that in other European countries. The EMR values in the neighborhood of 1% in males aged 40-64 that we observe for several Bulgarian regions are around or even in excess of many of the cited above IFR values for the whole population, and well in excess of most estimates for working age demographics in particular ^39^. Therefore, the potential impact of the SARS-CoV-2 virus for working age people may well have been underestimated previously.

## Methods

### Data Sources

All-cause mortality data for European countries and for NUTS-3 (Nomenclature of Territorial Units for Statistics) regions in Bulgaria was obtained from Eurostat ^40,41^. The data featured in these datasets is sex- and age-stratified, with age groups split in increments of 5 years.

Country-level population data was collected through Eurostat ^42^, and was further supplemented by population data from the United Nations’ UNdata Data Service ^43^. We further elaborate on this topic in the subsequent section on Potential Years of Life Lost (PYLL) and Working Years of Life Lost (WYLL) estimates.

The preliminary data from the most recent population census in Bulgaria was used for the analysis at the regional level in the country ^8^.

Life expectancy values at different ages were obtained from three separate sources. We acquire the full life tables for Bulgaria through the country’s National Statistical Institute ^44^. Abridged life tables for all European countries were obtained from the World Health Organization’s open data platform ^45^. This dataset is partitioned by age, in increments of 5 years. Abridged life tables for Bulgarian regions were created using regional mortality data for 2017– 2019 collected by Bulgaria’s National Statistical Institute ^44^ following the methodology of the ONS ^9^.

COVID-related mortality and testing data for Bulgaria was obtained from the Bulgaria’s Ministry of Health. The dataset, which covers the period from the beginning of the pandemic till March 2022, includes information about each infected individual’s age, gender, region, the date of their latest COVID-19 test, their status (infected, recovered, hospitalized, deceased), their hospitalization start and end dates, if any, whether they were taken into intensive care and whether they died of COVID-19.

## Data Availability

All datasets and associated code can be found at https://github.com/Mlad-en/Bulgaria_Regional_Mortality and https://github.com/Mlad-en/COV-BG.

## Excess mortality and P-scores

To calculate excess mortality across countries as well as across Bulgarian regions, we analyze the mortality observed between week 10 of 2020 and week 13 of 2022, and compare it to expected (baseline) mortality using the historical data for the five pre-pandemic years (2015–2019). The model we used is the Karlinsky–Kobak regression model ^4^:

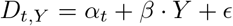

Where *D*_*t,Y*_ is the number of deaths observed in week or month *t* in year *Y, β* is a linear slope across years, and *α*_*t*_ are separate intercepts (fixed effects) for each week or month, and *ϵ* ∼ 𝒩(0, *σ*^2^) is Gaussian noise. The model prediction for a year *Y*, where *Y* = 2020, 2021 or 2022 is Expected Mortality 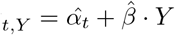.

We then establish a 95% confidence interval for the expected mortality. This range is used to calculate the excess mortality Δ_*t*_ for a week or a month *t* and a year *Y* as:

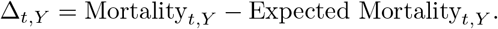

This calculation is done both as a sex- and age-stratified metric, as well as an aggregated total excess mortality for a year *Y*, which we denote by Δ_*Y*_. To normalize excess mortality across countries, we calculate excess mortality per total population. To do this, we use population data from Eurostat for 2020.

Set 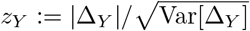, where Var[Δ_*Y*_] is computed in ^4^. If *z*_*Y*_ is significantly below 2 for a given country, we consider the excess mortality for this country to be not significantly different from zero. In the computations related to the years-of-life lost metrics considered in the paper, we excluded a few countries having both *z*_*Y*_ -values significantly below 2 (typically less than 1) for each age interval and wide confidence intervals that included 0 for the excess mortality associated with each of these age intervals.

Based on the excess mortality ranges we also compute a P-score value for each country/region. A P-score value is defined as the ratio or percentage of excess deaths over certain period relative to the expected deaths for the same period based on historical data from the years 2015–2019 (see ^46^). We calculate the P-score for a year *Y* as follows:

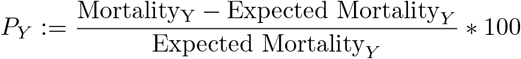

To calculate a total *P* -score we replace each term in in the right-hand side in the formula above by the corresponding summation over the three-year period considered in our analysis. We also calculate the ratio between excess mortality and official COVID-19-attributed mortality. Due to the demonstrably low testing in Bulgaria ^47^ and other countries, this allows us to estimate under-reported COVID-19 fatalities. We also use the total positive tests per region to compute a Case Fatality Ratio (CFR) which estimates the proportion of COVID-19 fatalities among confirmed cases.

### Potential Years of Life Lost (PYLL), Aged-Standardized Years of life lost Rate (ASYR), and Working Years of Life Lost (WYLL) estimates

Potential Years of Life Lost (PYLL) is a metric that estimates the burden of disease on a given population by looking at premature mortality. It is derived as the difference between a person’s age at the time died and the expected years of life for people at that age in a given country. As such, the metric attributes more weight to people that have died at a younger age.

We compute the PYLL across countries and Bulgarian regions by taking the positive all-cause excess mortality for all ages groups (in Eurostat they are aggregated at 5 year intervals). For the European countries considered in our paper we use the abridged life expectancy tables by the WHO (also aggregated at 5 year intervals) and for the Bulgarian regions we create abridged life expectancy tables following the ONS methodology ^9^ to calculate a total and average PYLL value for all countries and Bulgarian regions. To be more precise, for an age interval [*x, x* + 4] and sex *s* (if no sex is specified we assume it’s for both sexes) define by ED([*x, x* + 4], *s*) the excess deaths and by LE([*x, x* + 4], *s*) the life expectancy. Then the potential years of life lost are computed as

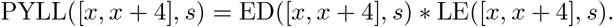

The total PYLL is computed by summing over all age intervals. In our computations we take into account the margin of error for each ED([*x, x* + 4], *s*).

A limitation on this approach is the upper-boundary aggregation value for the two datasets. The all-cause mortality dataset’s upper boundary is 90+, while the WHO’s abridged life tables only go up to the 85+ age bracket. To account for this, we attribute the life expectancy of the 85+ age group to the 85-89 mortality group. We have further excluded the 90+ mortality group from our analysis.

Finally, we standardize PYLL values across countries by dividing the total sum value by the population and normalizing it per 100,000 people:

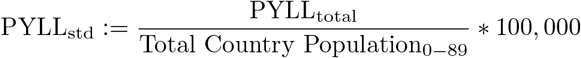

The data for country-level populations in Eurostat has a similar limitation in the upper boundary of the age distribution (a cut-off at 85+). To mitigate this limitation, we supplement the population data from Eurostat for ages 0-84 with population size data for the 85-89 age group from the UNdata Data Service.

To compare the impact of the pandemic across European populations and Bulgarian regions with different age structures we compute the Age-Standardized Years of Life Lost Rate (ASYR) ^48,49^. Let ([*x, x*+4], *s*) be an age interval for a sex *s* in a standard life expectancy table for a given population. Denote by *P* ([*x, x* + 4], *s*) the population size of ([*x, x* + 4], *s*). Define the PYLL rate for ([*x, x* + 4], *s*) as

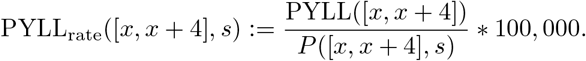

For the 2013 European Standard Population (ESP) denote by *W* ([*x, x* + 4], *s*) the weight of ([*x, x* + 4], *s*) in the standard population. Define

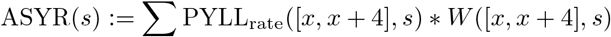

where the sum is taken over all age intervals. For a given population of sex *s* this measure is interpreted as the years of life lost per 100, 000 people (of sex *s*) if the population has the same age distribution as the ESP. We do the same for the Bulgarian regions using a standardized population for Bulgaria based on 2019 census estimates by the Bulgarian NSI. ASYR allows for comparison of the pandemic impact on EU countries and Bulgarian regions having different age distributions.

Finally, we derive total, average and total standardized WYLL value approximations. To accomplish this, we first assume people to be in the working age group if they are 15 to 64 years old, and thus exclude excess mortality for all age groups over 65. To calculate the remaining years of working life, we further assume a mean age for each age group, e.g. for the age interval 60 − 64 we assume a mean age at 62.5 years. This would leave this group with approximately 2.5 years until retirement. 95% CI for EMRs, P-scores and the values of all years of life lost functions can be found in our GitHub repository.

## Notes

### IRB and/or ethics committee approval

The Ethics Committee of IMI-BAS (Institute of Mathematics and Informatics, Bulgarian Academy of Sciences) gave ethical approval for this work.

## Author contributions

A.R, G.K.M. and M.M. conceived the project, analyzed the data and wrote the manuscript.

## Acknowledgments and Funding

The authors would like to acknowledge the help of the Bulgarian Ministry of Health and Information Services for providing us with raw data about COVID-19 mortality, hospitalizations and testing. A.R. would like to acknowledge the financial support of a “Petar Beron i NIE” fellowship [KP-06-D15-1] from the Bulgarian Science Fund.

## Supplementary Materials

### Supplementary Figures

**Supplementary Figure 1:**
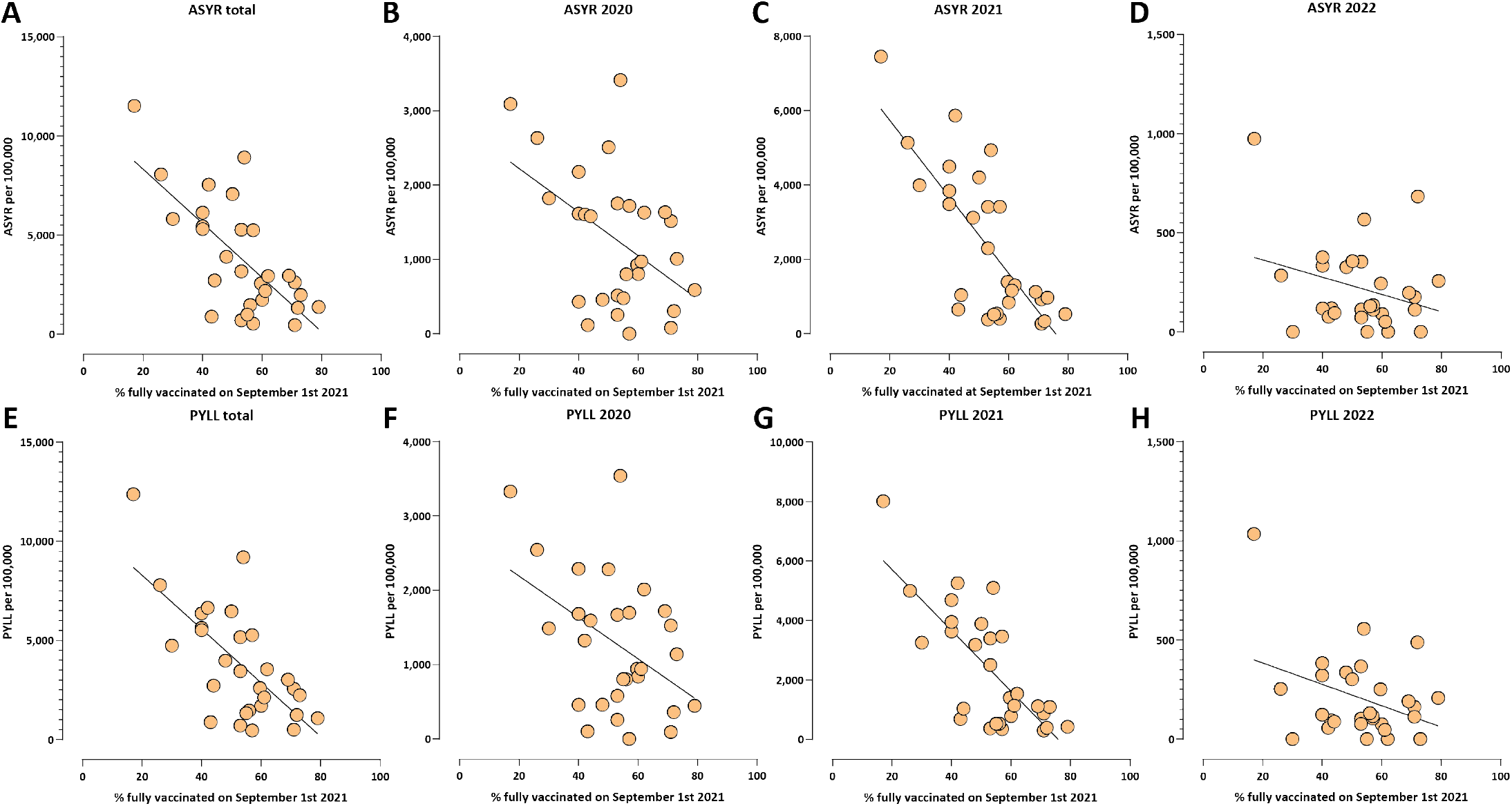
Correlation between rates of full vaccination and ASYR and PYLL excess mortality measures in European countries. (A) ASYR, total (Pearson *R*^2^ = 0.5, *p* ≤ 0.0001; Spearman *r* = −0.64, *p* = 0.0002); (B) ASYR, 2020 (Pearson *R*^2^ = 0.22, *p* = 0.0105; Spearman *r* = −0.36, *p* = 0.0549); (C) ASYR, 2021 (Pearson *R*^2^ = 0.57, *p* ≤ 0.0001; Spearman *r* = −0.69, *p* ≤ 0.0001); (D) ASYR, 2022 (Pearson *R*^2^ = 0.08, *p* = 0.1286; Spearman *r* = −0.2, *p* = 0.2982); (E) PYLL, total (Pearson *R*^2^ = 0.47, *p* ≤ 0.0001; Spearman *r* = −0.62, *p* = 0.0003); (F) PYLL, 2020 (Pearson *R*^2^ = 0.19, *p* = 0.0169; Spearman *r* = −0.32, *p* = 0.08); (G) PYLL, 2021 (Pearson *R*^2^ = 0.56, *p* ≤ 0.0001; Spearman *r* = −0.65, *p* = 0.0001); (H) PYLL, 2022 (Pearson *R*^2^ = 0.13, *p* = 0.0523; Spearman *r* = −0.19, *p* = 0.3106).

**Supplementary Figure 2:**
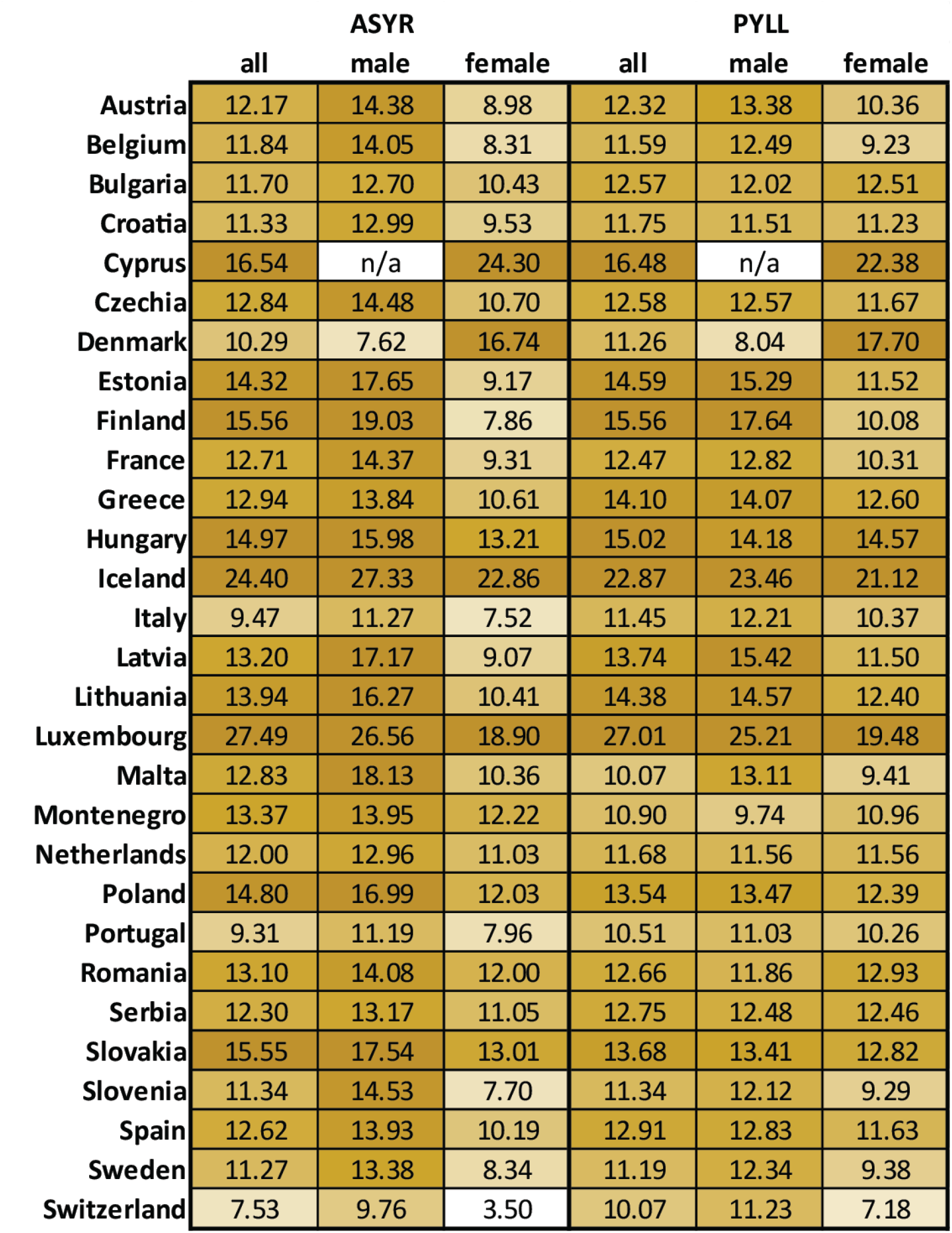
Year of life lost per excess death in European countries. Note that the very high values for a few countries (e.g. Cyprus, Iceland, Luxembourg, Finland, Malta) might be artifacts resulting from significantly insignificant excess deaths (z-score significantly below 2).

**Supplementary Figure 3:**
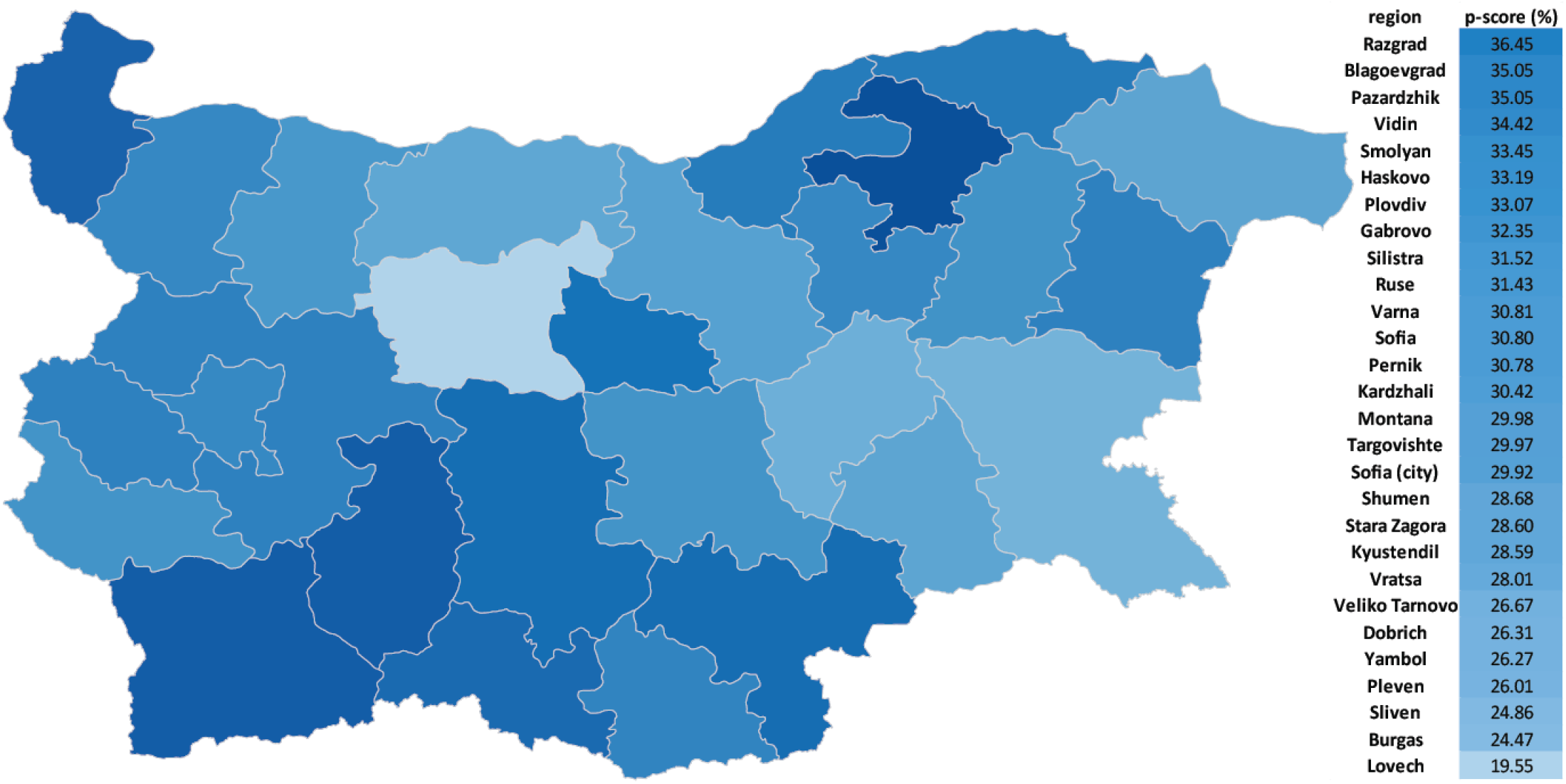
Excess mortality in Bulgarian regions (P-scores).

**Supplementary Figure 4:**
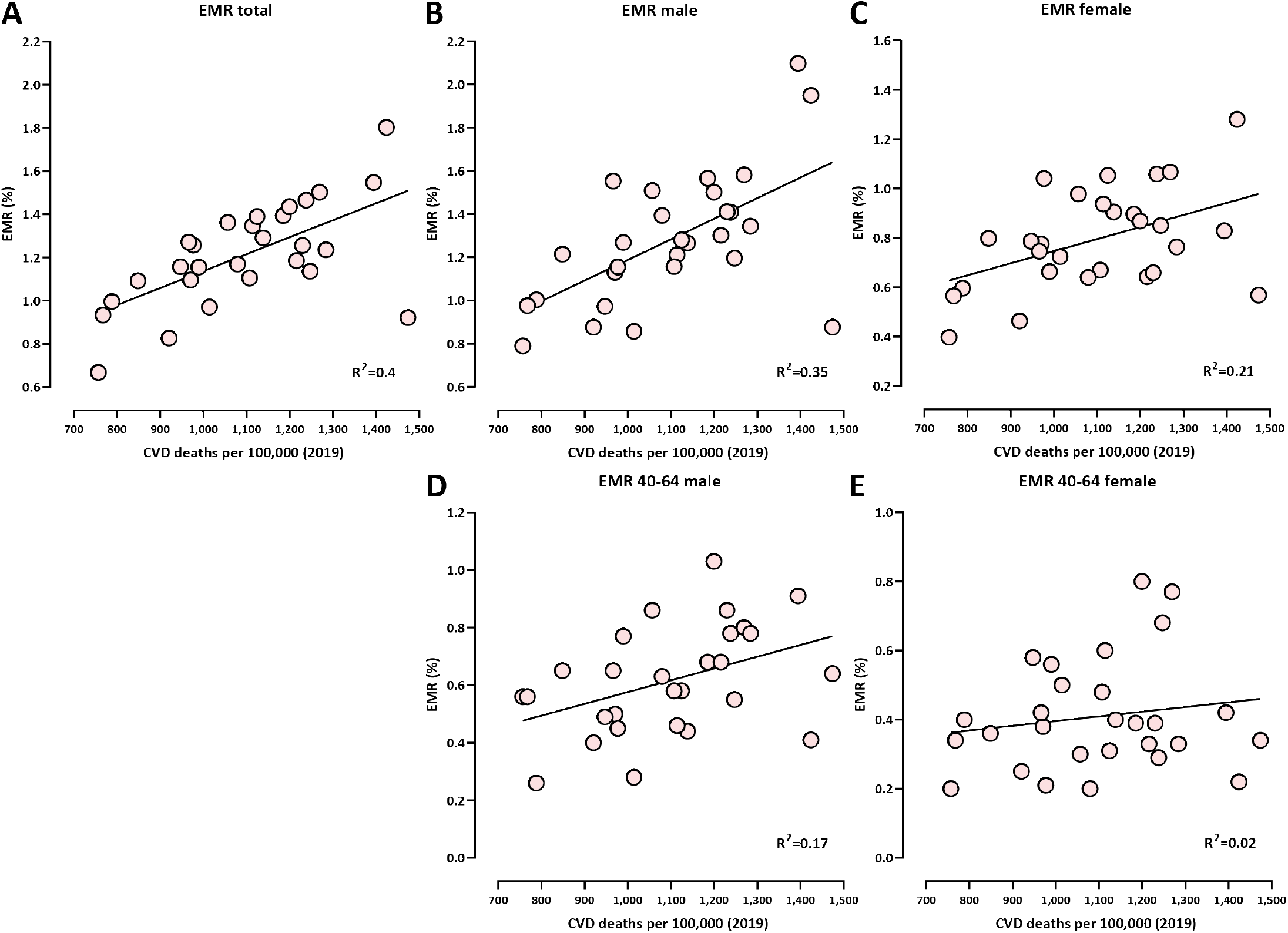
Correlation between cardiovascular disease (CVD) prevalence (measured as the deaths from CVD per 100,000 people in 2019) and COVID-related excess mortality in Bulgarian regions (as measured by EMR). Shown is the Pearson *R*^2^ correlation coefficient. (A) total (Pearson *R*^2^ = 0.4, *p* = 0.0003; Spearman *r* = 0.59, *p* = 0.0008); (B) males (Pearson *R*^2^ = 0.35, *p* = 0.0009; Spearman *r* = 0.57, *p* = 0.0014); (C) females (Pearson *R*^2^ = 0.21, *p* = 0.0129; Spearman *r* = 0.43, *p* = 0.0196); (D) males ages 40-64 (Pearson *R*^2^ = 0.17, *p* = 0.026; Spearman *r* = 0.43, *p* = 0.0214); (E) females ages 40-64 (Pearson *R*^2^ = 0.02, *p* = 0.41; Spearman *r* = 0.12, *p* = 0.51).

**Supplementary Figure 5:**
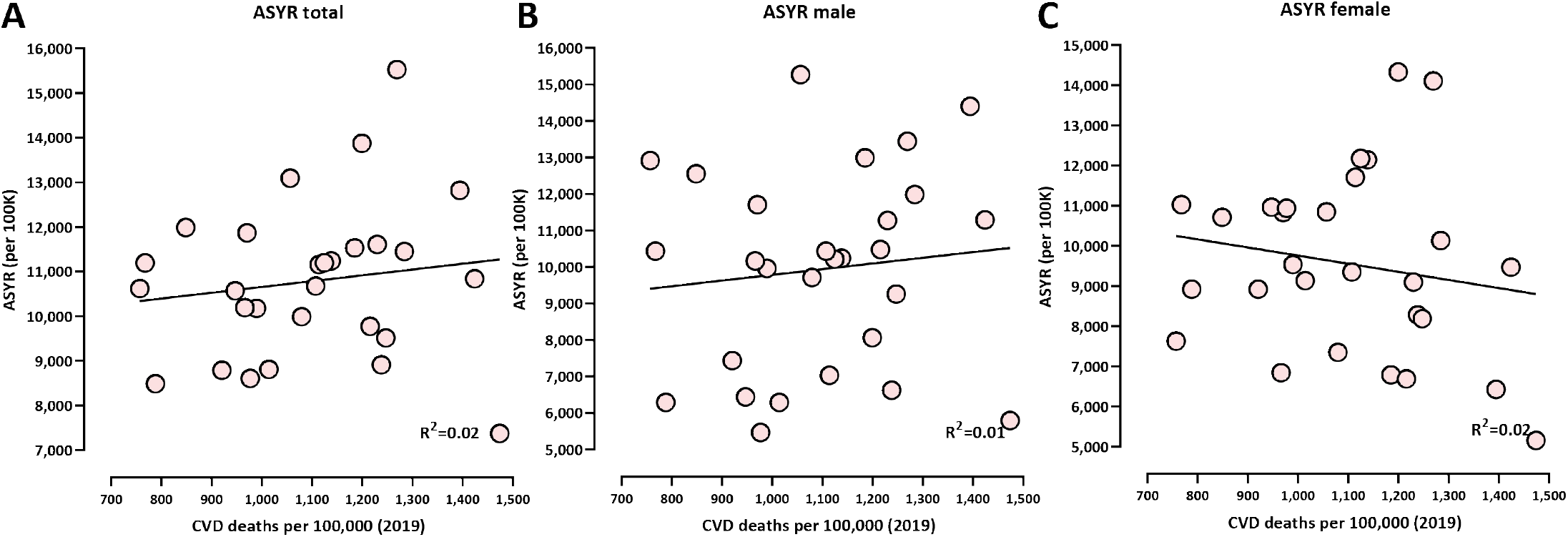
Correlation between cardiovascular disease (CVD) prevalence (measured as the deaths from CVD per 100,000 people in 2019) and COVID-related excess mortality in Bulgarian regions (as measured by ASYR). Shown is the Pearson *R*^2^ correlation coefficient. (A) total (Pearson *R*^2^ = 0.02, *p* = 0.47; Spearman *r* = 0.18, *p* = 0.35); (B) males (Pearson *R*^2^ = 0.01, *p* = 0.31; Spearman *r* = 0.15, *p* = 0.44); (C) females (Pearson *R*^2^ = 0.03, *p* = 0.79; Spearman *r* = −0.16, *p* = 0.40).

## References

1. Wang C, Horby PW, Hayden FG, Gao GF. 2020. A novel coronavirus outbreak of global health concerns. Lancet 395(10223):470–473.

2. Zhou P, Yang XL, Wang XG, Hu B, Zhang L, Zhang W, Si HR, Zhu Y, Li B, Huang CL, Chen HD, Chen J, Luo Y, Guo H, Jiang RD, Liu MQ, Chen Y, Shen XR, Wang X, Zheng XS, Zhao K, Chen QJ, Deng F, Liu LL, Yan B, Zhan FX, Wang YY, Xiao GF, Shi ZL. 2020. A pneumonia outbreak associated with a new coronavirus of probable bat origin. Nature 579(7798):270–273.

3. Huang C, Wang Y, Li X, Ren L, Zhao J, Hu Y, Zhang L, Fan G, Xu J, Gu X, Cheng Z, Yu T, Xia J, Wei Y, Wu W, Xie X, Yin W, Li H, Liu M, Xiao Y, Gao H, Guo L, Xie J, Wang G, Jiang R, Gao Z, Jin Q, Wang J, Cao B. 2020. Clinical features of patients infected with 2019 novel coronavirus in Wuhan, China. Lancet 395(10223):497–506.

4. Karlinsky A, Kobak D. 2021. The World Mortality Dataset: Tracking excess mortality across countries during the COVID-19 pandemic sElife 10:e69336

5. Pifarré I, Arolas H, Acosta E, López-Casasnovas G, Lo A, Nicodemo C, Riffe T, Myrskylä M. 2021. Years of life lost to COVID-19 in 81 countries. Sci Rep 11(1):3504.

6. Rangachev A, Marinov GK, Mladenov M. 2021. The demographic and geographic impact of the COVID pandemic in Bulgaria and Eastern Europe in 2020. Sci Rep 12(1):6333.

7. Ritchie H, Mathieu E, Rodés-Guirao L, Appel C, Giattino C, Ortiz-Ospina E, Hasell J, Macdonald B, Beltekian D, Roser M. 2020. Coronavirus Pandemic (COVID-19). Published online at OurWorldInData.org. Retrieved from: https://covid.ourworldindata.org/data/owid-covid-data.csv

8. Bulgarian National Statistical Institute. Preliminary assessment of the population of Bulgaria at September 7th 2021,

9. Office for National Statistics. 2021. Comparisons of all-cause mortality between European countries and regions: 2020. https://www.ons.gov.uk/peoplepopulationandcommunity/birthsdeathsandmarriages/deaths/articles/comparisonsofallcausemortalitybetweeneuropeancountriesandregions/2020.

10. Davies NG, Abbott S, Barnard RC, Jarvis CI, Kucharski AJ, Munday JD, Pearson CAB, Russell TW, Tully DC, Washburne AD, Wenseleers T, Gimma A, Waites W, Wong KLM, van Zandvoort K, Silverman JD; CMMID COVID-19 Working Group; COVID-19 Genomics UK (COG-UK) Consortium, Diaz-Ordaz K, Keogh R, Eggo RM, Funk S, Jit M, Atkins KE, Edmunds WJ. 2021. Estimated transmissibility and impact of SARS-CoV-2 lineage B.1.1.7 in England. Science 372(6538):eabg3055

11. Mlcochova P, Kemp SA, Dhar MS, Papa G, Meng B, Ferreira IATM, Datir R, Collier DA, Albecka A, Singh S, Pandey R, Brown J, Zhou J, Goonawardane N, Mishra S, Whittaker C, Mellan T, Marwal R, Datta M, Sengupta S, Ponnusamy K, Radhakrishnan VS, Abdullahi A, Charles O, Chattopadhyay P, Devi P, Caputo D, Peacock T, Wattal C, Goel N, Satwik A, Vaishya R, Agarwal M; Indian SARS-CoV-2 Genomics Consortium (INSACOG); Genotype to Phenotype Japan (G2P-Japan) Consortium; CITIID-NIHR BioResource COVID-19 Collaboration, Mavousian A, Lee JH, Bassi J, Silacci-Fegni C, Saliba C, Pinto D, Irie T, Yoshida I, Hamilton WL, Sato K, Bhatt S, Flaxman S, James LC, Corti D, Piccoli L, Barclay WS, Rakshit P, Agrawal A, Gupta RK. 2021. SARS-CoV-2 B.1.617.2 Delta variant replication and immune evasion. Nature 599(7883):114–119.

12. COVID-19 Forecasting Team. 2022. Variation in the COVID-19 infection-fatality ratio by age, time, and geography during the pre-vaccine era: a systematic analysis. Lancet 399(10334):1469–1488.

13. Murison KR, Grima AA, Simmons AE, Tuite AR, Fisman DN. 2022. Severity of SARS-CoV-2 Infection in Pregnancy in Ontario: A Matched Cohort Analysis. Clin Infect Dis ciac544.

14. Challen R, Brooks-Pollock E, Read JM, Dyson L, Tsaneva-Atanasova K, Danon L. 2021. Risk of mortality in patients infected with SARS-CoV-2 variant of concern 202012/1: matched cohort study. BMJ 372:n579

15. Grima AA, Murison KR, Simmons AE, Tuite AR, Fisman DN. 2022. Relative Virulence of SARS-CoV-2 Among Vaccinated and Unvaccinated Individuals Hospitalized with SARS-CoV-2. Clin Infect Dis ciac41

16. Bhopal SS, Bhopal R. 2020. Sex differential in COVID-19 mortality varies markedly by age. Lancet 396(10250):532–533.

17. Shu Y, McCauley J. 2017. GISAID: Global initiative on sharing all influenza data - from vision to reality. Euro Surveill 22(13):30494

18. Plante JA, Liu Y, Liu J, Xia H, Johnson BA, Lokugamage KG, Zhang X, Muruato AE, Zou J, Fontes-Garfias CR, Mirchandani D, Scharton D, Bilello JP, Ku Z, An Z, Kalveram B, Freiberg AN, Menachery VD, Xie X, Plante KS, Weaver SC, Shi PY. 2021. Spike mutation D614G alters SARS-CoV-2 fitness. Nature 592(7852):116–121.

19. Yurkovetskiy L, Wang X, Pascal KE, Tomkins-Tinch C, Nyalile TP, Wang Y, Baum A, Diehl WE, Dauphin A, Carbone C, Veinotte K, Egri SB, Schaffner SF, Lemieux JE, Munro JB, Rafique A, Barve A, Sabeti PC, Kyratsous CA, Dudkina NV, Shen K, Luban J. 2020. Structural and Functional Analysis of the D614G SARS-CoV-2 Spike Protein Variant. Cell 183(3):739–751.e8.

20. Korber B, Fischer WM, Gnanakaran S, Yoon H, Theiler J, Abfalterer W, Hengartner N, Giorgi EE, Bhattacharya T, Foley B, Hastie KM, Parker MD, Partridge DG, Evans CM, Freeman TM, de Silva TI; Sheffield COVID-19 Genomics Group, McDanal C, Perez LG, Tang H, Moon-Walker A, Whelan SP, LaBranche CC, Saphire EO, Montefiori DC. 2020. Tracking Changes in SARS-CoV-2 Spike: Evidence that D614G Increases Infectivity of the COVID-19 Virus. Cell 182(4):812–827.e19.

21. Viana R, Moyo S, Amoako DG, Tegally H, Scheepers C, Althaus CL, Anyaneji UJ, Bester PA, Boni MF, Chand M, Choga WT, Colquhoun R, Davids M, Deforche K, Doolabh D, du Plessis L, Engelbrecht S, Everatt J, Giandhari J, Giovanetti M, Hardie D, Hill V, Hsiao NY, Iranzadeh A, Ismail A, Joseph C, Joseph R, Koopile L, Kosakovsky Pond SL, Kraemer MUG, Kuate-Lere L, Laguda-Akingba O, Lesetedi-Mafoko O, Lessells RJ, Lockman S, Lucaci AG, Maharaj A, Mahlangu B, Maponga T, Mahlakwane K, Makatini Z, Marais G, Maruapula D, Masupu K, Matshaba M, Mayaphi S, Mbhele N, Mbulawa MB, Mendes A, Mlisana K, Mnguni A, Mohale T, Moir M, Moruisi K, Mosepele M, Motsatsi G, Motswaledi MS, Mphoyakgosi T, Msomi N, Mwangi PN, Naidoo Y, Ntuli N, Nyaga M, Olubayo L, Pillay S, Radibe B, Ramphal Y, Ramphal U, San JE, Scott L, Shapiro R, Singh L, Smith-Lawrence P, Stevens W, Strydom A, Subramoney K, Tebeila N, Tshiabuila D, Tsui J, van Wyk S, Weaver S, Wibmer CK, Wilkinson E, Wolter N, Zarebski AE, Zuze B, Goedhals D, Preiser W, Treurnicht F, Venter M, Williamson C, Pybus OG, Bhiman J, Glass A, Martin DP, Rambaut A, Gaseitsiwe S, von Gottberg A, de Oliveira T. 2022. Rapid epidemic expansion of the SARS-CoV-2 Omicron variant in southern Africa. Nature 603(7902):679–686.

22. Cele S, Jackson L, Khoury DS, Khan K, Moyo-Gwete T, Tegally H, San JE, Cromer D, Scheepers C, Amoako DG, Karim F, Bernstein M, Lustig G, Archary D, Smith M, Ganga Y, Jule Z, Reedoy K, Hwa SH, Giandhari J, Blackburn JM, Gosnell BI, Abdool Karim SS, Hanekom W; NGS-SA; COMMIT-KZN Team, von Gottberg A, Bhiman JN, Lessells RJ, Moosa MS, Davenport MP, de Oliveira T, Moore PL, Sigal A. 2022. Omicron extensively but incompletely escapes Pfizer BNT162b2 neutralization. Nature 602(7898):654–656.

23. Uraki R, Kiso M, Iida S, Imai M, Takashita E, Kuroda M, Halfmann PJ, Loeber S, Maemura T, Yamayoshi S, Fujisaki S, Wang Z, Ito M, Ujie M, Iwatsuki-Horimoto K, Furusawa Y, Wright R, Chong Z, Ozono S, Yasuhara A, Ueki H, Sakai-Tagawa Y, Li R, Liu Y, Larson D, Koga M, Tsutsumi T, Adachi E, Saito M, Yamamoto S, Hagihara M, Mitamura K, Sato T, Hojo M, Hattori SI, Maeda K, Valdez R; IASO study team, Okuda M, Murakami J, Duong C, Godbole S, Douek DC, Maeda K, Watanabe S, Gordon A, Ohmagari N, Yotsuyanagi H, Diamond MS, Hasegawa H, Mitsuya H, Suzuki T, Kawaoka Y. 2022. Characterization and antiviral susceptibility of SARS-CoV-2 Omicron BA.2. Nature 607(7917):119–127.

24. Yamasoba D, Kimura I, Nasser H, Morioka Y, Nao N, Ito J, Uriu K, Tsuda M, Zahradnik J, Shirakawa K, Suzuki R, Kishimoto M, Kosugi Y, Kobiyama K, Hara T, Toyoda M, Tanaka YL, Butlertanaka EP, Shimizu R, Ito H, Wang L, Oda Y, Orba Y, Sasaki M, Nagata K, Yoshimatsu K, Asakura H, Nagashima M, Sadamasu K, Yoshimura K, Kuramochi J, Seki M, Fujiki R, Kaneda A, Shimada T, Nakada TA, Sakao S, Suzuki T, Ueno T, Takaori-Kondo A, Ishii KJ, Schreiber G; Genotype to Phenotype Japan (G2P-Japan) Consortium, Sawa H, Saito A, Irie T, Tanaka S, Matsuno K, Fukuhara T, Ikeda T, Sato K. 2022. Vi-rological characteristics of the SARS-CoV-2 Omicron BA.2 spike. Cell 185(12):2103–2115.e19

25. Abdullah F, Myers J, Basu D, Tintinger G, Ueckermann V, Mathebula M, Ramlall R, Spoor S, de Villiers T, Van der Walt Z, Cloete J, Soma-Pillay P, Rheeder P, Paruk F, Engelbrecht A, Lalloo V, Myburg M, Kistan J, van Hougenhouck-Tulleken W, Boswell MT, Gray G, Welch R, Blumberg L, Jassat W. 2022. Decreased severity of disease during the first global omicron variant covid-19 outbreak in a large hospital in tshwane, south africa. Int J Infect Dis 116:38–42.

26. Maslo C, Friedland R, Toubkin M, Laubscher A, Akaloo T, Kama B. 2022. Characteristics and Outcomes of Hospitalized Patients in South Africa During the COVID-19 Omicron Wave Compared With Previous Waves. JAMA 327(6):583–584

27. Molenberghs G, Faes C, Verbeeck J, Deboosere P, Abrams S, Willem L, Aerts J, Theeten H, Devleesschauwer B, Bustos Sierra N, Renard F, Herzog S, Lusyne P, Van der Heyden J, Van Oyen H, Van Damme P, Hens N. 2022. COVID-19 mortality, excess mortality, deaths per million and infection fatality ratio, Belgium, 9 March 2020 to 28 June 2020. Euro Surveill 27(7):2002060

28. Pollán M, Pérez-Gómez B, Pastor-Barriuso R, Oteo J, Hernán MA, Pérez-Olmeda M, SanmartAñ JL, Fernández-García A, Cruz I, Fernández de Larrea N, Molina M, Rodríguez-Cabrera F, MartAñ M, Merino-Amador P, León Paniagua J, Muñoz-Montalvo JF, Blanco F, Yotti R; ENE-COVID Study Group. 2020. Prevalence of SARS-CoV-2 in Spain (ENE-COVID): a nationwide, population-based seroepidemiological study. Lancet 396(10250):535–544.

29. Kenyon C. 2020. COVID-19 Infection Fatality Rate Associated with Incidence-A Population-Level Analysis of 19 Spanish Autonomous Communities. Biology (Basel) 9(6):128

30. Ghisolfi S, Almøas I, Sandefur JC, von Carnap T, Heitner J, Bold T. 2020. Predicted COVID-19 fatality rates based on age, sex, comorbidities and health system capacity. BMJ Glob Health 5(9):e003094

31. Russell TW, Hellewell J, Jarvis CI, van Zandvoort K, Abbott S, Ratnayake R, Cmmid Covid-Working Group, Flasche S, Eggo RM, Edmunds WJ, Kucharski AJ. 2020. Estimating the infection and case fatality ratio for coronavirus disease (COVID-19) using age-adjusted data from the outbreak on the Diamond Princess cruise ship, February 2020. Euro Surveill 25(12):2000256.

32. Hauser A, Counotte MJ, Margossian CC, Konstantinoudis G, Low N, Althaus CL, Riou J. 2020. Estimation of SARS-CoV-2 mortality during the early stages of an epidemic: A modeling study in Hubei, China, and six regions in Europe. PLoS Med 17(7):e1003189

33. Meyerowitz-Katz G, Merone L. 2020. A systematic review and meta-analysis of published research data on COVID-19 infection fatality rates. Int J Infect Dis 101:138–148.

34. Thomas BS, Marks NA. 2021. Estimating the case fatality ratio for COVID-19 using a time-shifted distribution analysis. Epidemiol Infect 149:e197

35. Mizumoto K, Kagaya K, Chowell G. 2020. Early epidemiological assessment of the transmission potential and virulence of coronavirus disease 2019 (COVID-19) in Wuhan City, China, January-February, 2020. BMC Med 18(1):217.

36. Ioannidis JPA. 2021. Reconciling estimates of global spread and infection fatality rates of COVID-19: An overview of systematic evaluations. Eur J Clin Invest 51(5):e13554.

37. Bendavid E, Mulaney B, Sood N, Shah S, Bromley-Dulfano R, Lai C, Weissberg Z, Saavedra-Walker R, Tedrow J, Bogan A, Kupiec T, Eichner D, Gupta R, Ioannidis JPA, Bhattacharya J. 2021. COVID-19 antibody seroprevalence in Santa Clara County, California. Int J Epidemiol 50(2):410–419.

38. Marinov GK, Mladenov M, Rangachev A, Alexiev I. 2022. SARS-CoV-2 reinfections during the first three major COVID-19 waves in Bulgaria. medRxiv 2022.03.11.22271527.

39. Levin AT, Hanage WP, Owusu-Boaitey N, Cochran KB, Walsh SP, Meyerowitz-Katz G. 2020. Assessing the age specificity of infection fatality rates for COVID-19: systematic review, meta-analysis, and public policy implications. Eur J Epidemiol 35(12):1123–1138.

40. Eurostat. Deaths by week, sex, 5-year age group, https://ec.europa.eu/eurostat/databrowser/view/demo_r_mwk_05/default/table?lang=en.

41. Eurostat. Deaths by week, sex, 5-year age group and NUTS-3 region, https://ec.europa.eu/eurostat/databrowser/view/demo_r_mweek3/default/table?lang=en.

42. Eurostat. Population on 1 January by age group and sex region, https://ec.europa.eu/eurostat/databrowser/view/demo_pjangroup/default/table?lang=en.

43. UNdata Data Service. Population by age, sex and urban/rural residence, http://data.un.org/Data.aspx?d=POP&f=tableCode%3A22.

44. Bulgarian National Statistical Institute. Mortality and life expectancy by sex and place of residence, https://www.nsi.bg/en/content/6643/mortality-and-life-expectancy-sex-and-place-residence.

45. World Health Organization. Expectation of life at age X (Mortality and global health estimates, https://apps.who.int/gho/data/node.imr.LIFE_0000000035?lang=en.

46. Our World In Data. Excess mortality during the Coronavirus pandemic (COVID-19) https://ourworldindata.org/excess-mortality-covid#how-is-excess-mortality-measured.

47. Our World In Data. Total COVID-19 tests per 1,000 people, Our World in Data https://ourworldindata.org/grapher/full-list-cumulative-total-tests-per-thousand?time=2020-03-01..2020-12-31

48. Preedy V, Watson R (Eds.). 2010. Handbook of Disease Burdens and Quality of Life Measures. Springer.

49. Martinez R, Soliz P, Caixeta R, Ordunez P. 2019. Reflection on modern methods: years of life lost due to premature mortalityâ€”a versatile and comprehensive measure for monitoring non-communicable disease mortality. Int J Epidemiol.

